# Beyond Association: A Quantitative Analysis of the Infectious Burden in Alzheimer’s Disease

**DOI:** 10.64898/2025.12.24.25342962

**Authors:** Omer Bender, Or Shemesh, Daniel Z Bar

## Abstract

The etiology of Alzheimer’s disease (AD) remains a subject of intense investigation. While the 2024 Lancet Commission report attributes approximately 45% of global dementia cases to 14 modifiable risk factors, it notably excludes infectious agents due to debated causality. This exclusion contrasts with growing evidence that pathogens, specifically Herpes Simplex Virus Type 1 (HSV-1), *Porphyromonas gingivalis* and *Chlamydia pneumoniae*, may drive neuroinflammation, tau pathology and amyloidogenesis. This paper evaluates the Pathogen Hypothesis of AD through a quantitative framework. Synthesizing data from nationwide cohort studies, we apply a Population Attributable Fraction (PAF) model to estimate the disease burden driven by infectious agents. Our sensitivity analyses, utilizing E-values to quantify robustness against unmeasured confounding, suggest that infectious burden infectious burden may account for a plausibility range of **19% to 31%** from single-pathogen models, which expands to **31% to 52%** in joint models of sporadic AD cases. We explicitly model the heterogeneity between cohorts and the synergistic interaction with the APOE ε4 allele. Furthermore, we address recent failures in antimicrobial clinical trials (VALAD, GAIN), arguing that these outcomes reflect a need for precision biomarker stratification rather than a refutation of the hypothesis. These findings argue for the integration of precision pathogen suppression into dementia prevention protocols.

## 1. Introduction

Alzheimer’s disease (AD) is a devastating, progressive neurodegenerative disorder and the most common cause of dementia [1,2]. For over three decades, the prevailing “Amyloid Cascade Hypothesis” posited that the accumulation of amyloid-beta (Aβ) plaques was the primary inciting event, triggering a cascade of neurodegeneration [3]. However, the modest clinical benefits of Aβ-clearing therapeutics have prompted a significant shift in focus toward other core pathological hallmarks. Chief among these are the accumulation of hyperphosphorylated tau (p-tau) into neurofibrillary tangles (NFTs) and chronic neuro-inflammation [2]. The anatomical spread of NFTs correlates more closely with the severity of cognitive decline [4,5]. Chronic neuro-inflammation, driven by activated glial cells, accelerates both Aβ aggregation and tau hyperphosphorylation [6].

This stagnation of the amyloid paradigm has redirected research efforts to better understand upstream factors like infectious mechanisms. Studies have found elevated Herpes Simplex Virus Type 1 (HSV-1) protein expression in AD brains [7–10]. This expression strongly colocalizes with p-tau, but not with Aβ plaques or oligomers [11]. This evidence suggests a novel function for tau phosphorylation, demonstrating that it acts as an innate immune response to HSV-1 infection [11]. This response is driven by the cGAS-STING pathway, which notably reduces viral protein expression and preserves neuronal viability. Collectively, these findings argue that inflammatory and infectious mechanisms may be the key initiators of both amyloidosis and tauopathy.

The “Antimicrobial Protection Hypothesis” proposes that amyloid-beta (Aβ) is not a metabolic waste product but a highly conserved antimicrobial peptide (AMP) of the innate immune system [12,13]. In this model, Aβ fibrillizes to entrap invading pathogens, which protects the brain from acute infection. Supporting this, studies have shown that HSV-1 infection can induce Aβ production [14,15]. However, in the aging brain, a compromised immune system, or immunosenescence, allows for the chronic reactivation of latent viruses such as HSV-1 or the persistent infiltration of bacteria like *Porphyromonas gingivalis [16–18]*. This chronic immune challenge drives the sustained production of “protective” amyloid, which eventually becomes neurotoxic, ultimately leading to synaptic loss and neurodegeneration.

Early investigations by Balin et al. established a strong association between Chlamydia pneumoniae and Alzheimer’s disease (AD), detecting the pathogen in 90% (17/19) of post-mortem AD brains compared to only 5% (1/19) of controls [19]. Subsequent immunohistochemical analyses have further characterized the cellular tropism of the infection, revealing that approximately 20% of neurons in affected AD brain regions harbor the bacterium, alongside significant infection in microglia and astrocytes [20,21]. A recent large-scale post-mortem analysis (n=95) corroborates these findings, reporting a 2.9- to 4.1-fold increase in C. pneumoniae inclusions in AD retinas and brain tissue, respectively, compared to controls [22]. Notably, this study utilized machine learning models to demonstrate that retinal C. pneumoniae burden, in combination with Aβ and NLRP3 levels, could predict AD diagnosis with high accuracy.

Epidemiological data reinforces the pathological findings. A nationwide cohort study from Taiwan (n=6,628) demonstrated that patients hospitalized for *C. pneumoniae* pneumonia exhibited a significantly increased risk of developing AD (HR = 1.599, 95% CI: 1.284–1.971), with the risk persisting over an 8-year follow-up [23]. Meta-analytic pooling of serological and post-mortem data yields an even stronger association, with an Odds Ratio (OR) of 5.66 (95% CI: 1.83–17.51) [24].

Importantly, infectious risk may be “polymicrobial”: cumulative and/or interacting pathogen burdens have been repeatedly studied (serology- and cohort-based), motivating models in which multiple microbes jointly shape neuroinflammation and cognitive trajectories rather than acting in isolation [25]. A biologically plausible oral–neurotropic synergy is emerging for HSV-1 and P. gingivalis: herpesvirus–bacteria coinfection is a recognized driver framework in periodontitis, and (mechanistically) the P. gingivalis lysine-gingipain Kgp can disable interferon signaling, increase HSV-1 infection/spread in human gingival models, and promote HSV-1 reactivation in neuronal cells [26].

C. pneumoniae also has an oral foothold relevant to this “gateway” concept—being detected more frequently in subgingival plaque from periodontitis sites (where classic periodontal pathogens such as P. gingivalis are also present), and clinical isolates have been recovered from gingival crevicular fluid—supporting a plausible route for repeated exposure alongside periodontal dysbiosis [27].

Although direct HSV-1 - Chlamydia pneumoniae co-infection studies in CNS cells remain limited, in vitro evidence indicates that the two pathogens converge on key Aβ generation/clearance pathways: C. pneumoniae infection of human astrocytes increases BACE1 and PSEN1 expression/activity while reducing ADAM10 activity, shifting APP processing toward a pro-amyloidogenic route; HSV-1 infection in neurons drives APP cleavage generating AICD that binds the neprilysin (NEP) promoter and dynamically modulates NEP expression and enzymatic activity [28,29].

Furthermore, the brain’s defense mechanism extends beyond Aβ, as phosphorylated tau (p-tau) has been demonstrated to serve as an innate immune response to HSV-1 infection, reducing viral protein expression and significantly decreasing post-infection neuronal death [11,30,31]. This suggests that p-tau is a critical innate immune anti-herpetic factor, highlighting the complex role of both Aβ and p-tau in the brain’s response to infection.

Despite biological plausibility, current public health models exclude infectious agents from modifiable risk factors. This paper aims to bridge the gap between hazard ratios and public health impact by estimating the Population Attributable Fraction (PAF) of specific pathogens. We provide a heuristic comparison to the Lancet Commission’s 45%, to contextualize its magnitude.

## 2. Methods

### 2.1 Population Attributable Fraction (PAF) Framework

To estimate the proportion of sporadic Alzheimer’s disease (AD) cases plausibly attributable to infectious pathogens, we employed Levin’s formula for the Population Attributable Fraction (PAF) [32]. The formula is defined as:

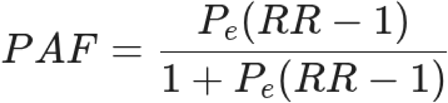

Where **P**_e_ represents the prevalence of the exposure in the population and **RR** represents the Relative Risk.

### 2.2 Assumptions and Causal Inference

We acknowledge that Levin’s formula assumes the input RR reflects a causal relationship and that the exposure is amenable to intervention. As our inputs are derived from observational epidemiology, these estimates are subject to confounding, selection bias, and reverse causation.

#### Estimand Harmonization

Where Hazard Ratios (HR) from longitudinal cohorts were available (e.g., for HSV-1), we utilized them as the most robust proxy for RR. Where only Odds Ratios (OR) were available, we treated them as approximations of RR, noting that this may overestimate the PAF if the outcome is common.

#### Sensitivity Analysis (E-Values)

To address unmeasured confounding, we calculated E-values for key inputs [33]. The E-value represents the minimum strength of association an unmeasured confounder would need to have with both the exposure and the outcome to explain away the observed risk. The formula used is:

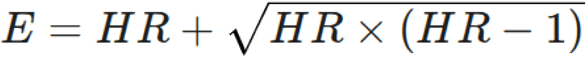

### 2.3 Addressing Joint Exposure and Additivity

#### Calculation of Total Pathogen Burden

Simple summation of individual pathogen PAFs leads to overestimation because exposures (e.g., HSV-1, *P. gingivalis*, and *C. pneumoniae*) are not mutually exclusive and likely co-occur in the same individuals. To estimate the combined burden of these pathogens, we avoided simple addition and instead calculated theoretical bounds based on the degree of biological overlap.

● **Lower Bound (Total Overlap/Redundancy)**: This scenario assumes that pathogen exposures are perfectly correlated or that a single dominant pathogen drives the pathology in co-infected individuals. The joint burden is therefore equal to the single strongest risk factor among the group:

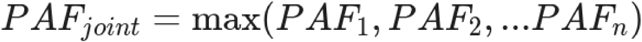
● **Theoretical Upper Bound (Independence)**: This scenario assumes that pathogen exposures occur independently and contribute additively to the risk (minus the intersection). We calculated this using the inclusion-exclusion principle:

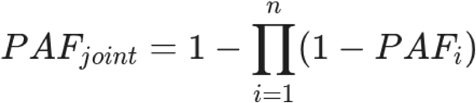

#### Exclusion of Etiological Estimates

For these joint calculations, we utilized only the conservative PAF estimates derived from systemic exposure (serology/longitudinal diagnoses) rather than the etiological estimates derived from CNS invasion. We excluded the CNS invasion estimates because their high individual values (∼80–90%) reflect a proximal stage of disease where causality is nearly absolute. This decision may need to be reconsidered if new large-scale studies reproduce the CNS invasion results.

## 3. Results: Pathogen-Specific Burden Estimates

### 3.1 Herpes Simplex Virus Type 1 (HSV-1)

HSV-1 is a neurotropic virus known to establish a lifelong latent infection primarily in the trigeminal ganglia. Quantifying the precise burden of this pathogen has been a key focus of the infectious hypothesis. Evidence from a landmark Taiwanese cohort study found an adjusted HR of 2.56 [33,34]. However, relying on medical coding creates specificity bias. Using a conservative “Clinical Reactivation” prevalence of 10% (P_e_ = 0.10) and the adjusted HR of 2.56, the specific PAF is **13.5%**.

#### Sensitivity Analysis (E-Value)

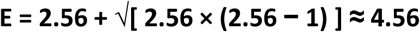

The E-value is **4.56**. This implies that an unmeasured confounder (e.g., a specific genetic vulnerability beyond APOE) would need to be associated with both HSV-1 reactivation and dementia by a risk ratio of at least 4.56 to explain away this association entirely. This suggests the association is moderately robust to confounding.

### 3.2 Chronic Periodontitis and *Porphyromonas gingivalis*

*P. gingivalis*, the main bacterium responsible for chronic periodontitis, produces toxic proteases called gingipains, which have been found in >90% of AD brains in some series, though replication varies [18]. Quantifying this risk, long-term chronic periodontitis (lasting >10 years) is associated with a significant adjusted Hazard Ratio (HR) of 1.71 (95% CI: 1.15–2.53) for AD [35]. Based on the PAF model, and using an exposure prevalence of 34% (for moderate-to-severe disease in the elderly), chronic periodontitis is estimated to contribute **19.4%** of sporadic AD cases.

#### Sensitivity Analysis (E-Value)

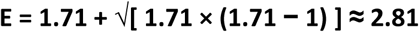

The E-value is 2.81. While lower than HSV-1, this indicates that a confounder would still need a substantial association (>2.8-fold) with both oral health and dementia to nullify the finding.

#### Etiological Estimate (CNS Invasion)

While chronic periodontitis serves as a proxy for systemic exposure, quantifying the risk based on direct CNS invasion yields a significantly higher burden. Post-mortem analysis by Dominy et al. [18] identified gingipains (toxic proteases) in 91–96% of AD brains compared to 39–52% of age-matched controls, depending on the specific protease targeted (Kgp vs. RgpB). Using these prevalence ranges, we estimated the etiological burden. Weighting these frequencies against AD prevalence suggests that approximately **45% to 56%** of the elderly population harbors *P. gingivalis* in the brain. The derived Relative Risk (RR) for those with CNS invasion ranges from **7.8** (conservative, based on Kgp) to **28.8** (upper-limit, based on RgpB). Consequently, the fraction of cases attributable to CNS invasion is estimated between **79% and 93%**.

#### Sensitivity Analysis (E-Value)

The E-value for the lower bound of this etiological estimate (RR = 7.82) is 15.12. This magnitude implies that for an unmeasured confounder to explain away the association between brain gingipains and Alzheimer’s, it would need to increase the risk of both by more than 15-fold—a degree of confounding that is virtually implausible for standard demographic or lifestyle factors.

### 3.3 The “Vaccine Signal”: Heterogeneity and Hypothesis Generation

Meta-analyses consistently show that specific adult vaccinations—particularly against Herpes Zoster (Shingles)—reduce the risk of dementia by approximately 25–40% (RR ≈ 0.70), with “natural experiments” more clear of user bias providing similar results [36–39]. This offers a potential “top-down” validation of the infectious hypothesis, suggesting that suppressing neurotropic pathogens can alter the disease trajectory.

However, this signal is complex and often obscured in broad aggregate analyses. As noted by critics, some large population-based studies have reported no overall protective effect, or even a paradoxical *increased* risk of dementia associated with common vaccines like influenza (OR 1.39) [40]. We argue that these “deleterious” findings are likely spurious artifacts of reverse causation and detection bias: individuals in the prodromal stages of dementia interact more frequently with the healthcare system, leading to higher vaccination rates than the general population [41]. Crucially, when these studies are corrected using a 10-year lag period or active comparators (e.g., cancer screening), the apparent “risk” dissipates [40].

When disaggregated, the signal for specific anti-herpetic interventions remains robust. The protective effect of Shingles vaccination (targeting VZV, a known trigger for HSV-1 reactivation) appears more consistent than that of generic respiratory vaccinations [36,40].

#### Sensitivity Analysis

The E-value for the aggregate vaccine RR of 0.70 is 2.21. This relatively low value suggests that Healthy User Bias—where conscientious individuals are both more likely to vaccinate and less likely to develop dementia—could plausibly explain much of the general vaccine signal. However, bias alone struggles to explain the specificity of the protection seen with Zoster vaccines compared to influenza or pneumococcal vaccines. Therefore, while we treat this data as hypothesis-generating rather than definitive validation, the “Zoster Signal” warrants prioritized investigation as a specific, pathogen-suppressing intervention.

### 3.4 Association of *Chlamydia pneumoniae* with Alzheimer’s Disease

Based on HR and OR data, we estimated the Population Attributable Fraction (PAF) for *C. pneumoniae* in AD.

● **Conservative Estimate:** Assuming a widespread seroprevalence of ∼75% in the elderly and the conservative Hazard Ratio of 1.6 from longitudinal data, the PAF is estimated at **∼31%**.
● **Etiological Estimate**: If exposure is defined strictly by CNS invasion (present in ∼85% of AD cases vs. ∼5% of controls), the PAF rises to ∼83% (reflecting that while systemic exposure is common, CNS invasion is estimated to affect only ∼14% of the elderly population but carries significantly higher risk).

This wide range reflects the uncertainty regarding whether systemic exposure or specific neuro-invasion is the primary driver of risk. Additional caution is required due to the variance in *C. pneumoniae* detection in AD brains (see discussion).

#### Sensitivity Analysis (E-Value)

Given the disparity between the longitudinal and meta-analytic risk estimates, we calculated E-values for both scenarios. For the conservative estimate based on the Taiwan cohort (HR = 1.599), the E-value is **2.58**. This indicates that an unmeasured confounder would need to increase the risk of both *C. pneumoniae* pneumonia and dementia by approximately 2.6-fold to explain away the observed association. This suggests moderate robustness, similar to that of chronic periodontitis.

However, for the etiological estimate derived from the meta-analysis (OR = 5.66), the E-value rises to **10.79**. This extremely high value implies that if the strong associations reported in pathological and serological meta-analyses are accurate, they are virtually impossible to explain via unmeasured confounding (e.g., smoking or socioeconomic status). The “robustness” here is limited not by confounding, but by the heterogeneity of the underlying detection methods discussed below.

#### Effect of Antimicrobial Intervention

Crucially, the Taiwan cohort provided evidence for the reversibility of this risk. Treatment with macrolides or fluoroquinolones for >15 days was associated with a significantly decreased risk of subsequent AD [23]. Fluoroquinolones, which possess superior blood-brain barrier penetrance compared to macrolides, showed a particularly robust protective effect, suggesting that central eradication of the pathogen may be necessary for therapeutic benefit.

## 4. Discussion

### Mechanisms of Pathogenesis

The data support a model where *C. pneumoniae* acts as a driver of AD pathology rather than a passive bystander. Murine models have demonstrated that the bacterium can rapidly bypass the blood-brain barrier via the olfactory and trigeminal nerves, seeding the CNS within 72 hours of intranasal inoculation [42–44]. Once in the brain, the infection appears to directly promote amyloidogenesis; in vivo studies show that *C. pneumoniae* infection alters APP processing via BACE1 and γ-secretase dysregulation, leading to the deposition of Aβ1-42 plaques. Furthermore, recent evidence highlights the activation of the NLRP3 inflammasome as a critical downstream effector. The correlation between *C. pneumoniae* burden and cleaved gasdermin D/caspase-3 levels suggests that the infection triggers pyroptotic and apoptotic cell death pathways, contributing to the neurodegeneration observed in the infected neuronal population.

Unlike C. pneumoniae, where controls are rarely infected, P. gingivalis is present in a significant proportion (∼39–52%) of cognitively normal elderly. This indicates that mere “presence” is not the sole determinant of pathology. Rather, the bacterial load—which was significantly higher in AD brains — and the host’s inflammatory threshold likely determine the transition from colonization to neurodegeneration. This reinforces the “Infectious Burden” model, where the quantity of CNS invasion, rather than binary exposure, drives the disease.

Co-infection provides a coherent “amplifier” mechanism across compartments. In the oral compartment, *P. gingivalis* can functionally weaken antiviral defenses via Kgp-mediated disruption of interferon signaling and thereby increase HSV-1 infection and propagation; notably, it has also been observed to promote HSV-1 reactivation in neuronal cells. This supports a mechanistic bridge by which chronic periodontitis could raise the probability (or frequency) of HSV-1 reactivation and downstream neuroinflammatory cascades [45].

In parallel, glial infection biology supports convergence on amyloidogenic processing: *C. pneumoniae* infection of human astrocytic cells shifts APP processing toward a more amyloidogenic profile, including increased BACE1 and PSEN1 and reduced ADAM10 activity. In addition, HSV-1 infection has been shown to modulate neprilysin (NEP) expression and activity through APP intracellular domain–related mechanisms, providing a plausible route by which concurrent or sequential infections could bias the system toward greater Aβ generation and reduced clearance [28].

Crucially, the risk associated with HSV-1 is not uniformly distributed. Research strongly indicates a synergistic interaction with the APOE ε4 allele [46]. In this model, HSV-1 confers the highest risk in APOE ε4 carriers, where the virus acts as the “match” that initiates the process, while the allele acts as the “gasoline,” impeding viral clearance.

### Limitations and Heterogeneity of Evidence

Despite compelling mechanistic and epidemiological data, the hypothesis faces significant challenges regarding reproducibility. While Balin et al. and recent studies report high detection rates, other groups have failed to replicate these findings, reporting low or absent *C. pneumoniae* DNA in AD brains [47–49]. These discrepancies likely stem from methodological variability, particularly the difficulty in detecting “aberrant bodies”—the persistent, non-replicative form of the bacterium that resists standard culture and PCR detection methods. Furthermore, the ubiquity of seropositivity in the elderly (>75%) [50] contrasts with the lower prevalence of AD (∼11%), creating a “ubiquity paradox.” This suggests that infection alone is insufficient to cause disease; rather, host-specific factors—such as APOE ε4 genotype, which facilitates bacterial attachment to glia, or olfactory barrier integrity—likely act as critical bottlenecks determining whether a common respiratory infection progresses to a chronic neurodegenerative condition. Consequently, while the estimated PAF suggests a massive potential impact, it must be interpreted with caution given the small sample sizes of the initial pathological cohorts and the high variability in risk estimates across studies.

For co-infection, the strongest evidence remains largely pairwise and tends to be compartment-specific, drawing from oral tissue models, glial cell systems, and serologic infection-burden studies. Large human neuropathology cohorts have not yet provided definitive demonstrations of simultaneous brain infection by HSV-1, *P. gingivalis*, and *C. pneumoniae* in the same individuals. Accordingly, co-infection is best framed as a mechanistically supported interaction hypothesis that may help explain heterogeneity across studies, rather than as established human neuropathology [45].

### 4.1 Synthesis of Burden and Joint Limits

To quantify the aggregate impact of a “polymicrobial” burden, we modeled the joint Population Attributable Fraction (PAF) for pairwise and triple combinations of the three pathogens. Using the conservative estimates—HSV-1 (13.5%), Chronic Periodontitis (19.4%), and *C. pneumoniae* (31%)—we derived the following plausibility ranges:

#### Pairwise Estimates

● **HSV-1 and *P. gingivalis*:** The joint burden lies between a lower bound of **19.4%** (Total Overlap) and an upper bound of **30.3%** (Independence).
● **HSV-1 and *C. pneumoniae*:** This pairing, relevant for their convergence on glial Aβ clearance pathways, yields a joint burden between **31%** and **40.3%**.
● ***P. gingivalis* and *C. pneumoniae*:** For this bacterial pairing, the burden is estimated between **31%** and **44.4%**.

#### Combined Triple Burden

When modeling the simultaneous contribution of all three pathogens (HSV-1 + *P. gingivalis* + *C. pneumoniae*), the Total Infectious Burden is estimated to account for a lower Bound of 31% (driven by the dominant contribution of *C. pneumoniae*) and an upper Bound of 51.9% (assuming independent contribution).

#### Synthesis

These calculations suggest that even under conservative assumptions—ignoring the high-risk “CNS invasion” subsets—the cumulative infectious burden plausibly accounts for **31% to 52%** of sporadic Alzheimer’s disease cases. This magnitude places the “infectious composite” risk on par with, or potentially exceeding, the combined weight of traditional vascular risk factors (e.g., hypertension, obesity, and diabetes) often cited in prevention models.

Notably, mechanistic co-infection undermines the assumption that exposures act independently. If periodontitis increases susceptibility to HSV-1 infection or promotes viral reactivation, then the joint effect is more appropriately modeled as conditionally dependent rather than independent. In that case, estimating combined impact requires joint-prevalence information or explicit interaction terms, rather than assuming a simple multiplicative relationship [45].

### 4.2 Reconciling the VALAD and GAINClinical Trials

A critical challenge to the infectious hypothesis is the translation of observational signals into successful interventions. While observational data suggests protective effects of antivirals, recent randomized controlled trials have failed to meet their primary endpoints.

#### VALAD Trial

Recent reports indicate that Valacyclovir did not significantly slow cognitive decline in patients with established AD [51–53].

#### GAIN Trial (COR388)

This study (NCT03823404), targeting gingipains with the inhibitor atuzaginstat, also missed its primary endpoint in the broad AD population [54,55].

These failures mirror the history of anti-amyloid trials: intervention may be occurring too late in the disease cascade, or in the wrong patients. Treating a general AD population with a specific antimicrobial agent will inevitably fail if a significant proportion of the cohort lacks the specific target pathogen, or if the “infectious ignition” occurred decades prior. The lack of efficacy in these trials likely reflects limitations of a “one-size-fits-all” approach rather than a fundamental refutation of the pathogen hypothesis.

### 4.3 Future Directions: Precision Medicine

To move from a broad “plausible range” toward true precision public health, future research must pivot decisively toward a precision medicine framework. Central to this shift is biomarker-based stratification. Clinical trials should require objective evidence of active pathogenic burden—such as elevated cerebrospinal fluid viral load or high-titer *P. gingivalis* markers, to ensure that therapeutic interventions are tested in biologically relevant patient populations. Matching the right patient to the right intervention is essential; consistent with this principle, post hoc analyses of the GAIN trial revealed that efficacy was confined to the subset of patients in whom *P. gingivalis* DNA was detectable.

Equally important is the timing of intervention. Antiviral or antimicrobial suppression may need to be initiated during the prodromal phase of disease, before downstream neuroinflammatory and neurodegenerative cascades become self-sustaining and less responsive to pathogen-directed therapies.

Finally, a co-infection framework argues against single-pathogen stratification in favor of multiplex pathogen profiling. Such an approach would incorporate markers of HSV activity or reactivation, measures of gingipain or Kgp exposure, and indicators of *C. pneumoniae* burden. Because interactions among pathogens can shape host immune responses and disease trajectories, cohorts classified as pathogen-positive may remain biologically heterogeneous unless co-pathogens are measured and explicitly modeled. Embracing this complexity will be critical for translating infectious hypotheses of neurodegeneration into actionable, precision-guided interventions.

## 5. Conclusion

Our quantitative analysis, supported by sensitivity modeling, suggests that pathogens plausibly contribute to approximately one-fifth to one-third of sporadic Alzheimer’s cases. While precise attribution is limited by the need for better biomarkers and formal overlap modeling, the convergence of epidemiological and mechanistic data is compelling. Public health strategies should urgently expand to include validatory biomarker research and precision trials as standard neuroprotection.

## Acknowledgments

We thank the Bar lab members for comments and suggestions. This work was supported by the Israel Science Foundation (grants 654/20 and 632/20 to DZB).

## Data Availability

Not applicable.

## Ethical Committee approval and Patient Consent

Not applicable.

